# Determinants of cholera fatalities in Malawi: A case-control study of patient-level and clinical management factors in the 2022-23 outbreak

**DOI:** 10.1101/2025.02.07.25321858

**Authors:** Ronald Chitatanga, Alex Thawani, Hope Chadwala, Amon Chirwa, Collins Mitambo

## Abstract

Malawi experienced its deadliest cholera outbreak in 2022, reporting over 50,000 cases and more than 1,700 deaths, resulting in a case fatality rate of 3.1%. The outbreak was exacerbated by the devastation caused by Tropical Cyclones Freddy, Ana, and Gombe, which strained healthcare resources. Despite the severity of the outbreak, no real-time evaluations of patient-level risk factors influencing cholera mortality among hospitalized patients were conducted. This study characterizes patient-level factors and treatment practices associated with cholera mortality in Malawi. A multisite case-control study was conducted in August 2023 across four cholera treatment units. A retrospective review of 174 medical records (confirmed by rapid antigen test or stool culture) was performed by a team led by trained medical officers. Cases (deceased) and controls (survivors) were matched based on age and district of diagnosis. Conditional logistic regression was used to assess associations between patient characteristics and mortality. Odds ratios (ORs) with 95% confidence intervals (CIs) were calculated for all predictors, with statistical significance set at p < 0.05. The strongest predictor of mortality was inadequate intravenous (IV) fluid management, with 95% of deceased patients classified as inadequately managed. Inadequate fluid management was associated with significantly higher odds of mortality (OR = 117, 95% CI: 14.3–959, p < 0.001). This study highlights critical gaps in adherence to cholera treatment protocols in Malawi’s treatment units, emphasizing the need for timely and adequate IV fluid management to reduce cholera-related deaths.

## INTRODUCTION

Between 1990 and 2019, cholera was responsible for an estimated three million global deaths (1). These fatalities ranged from 83,045 in 1990 to 117,167 in 2019, demonstrating an overall increasing trend in mortality. Alarmingly, the African Region accounted for the highest number of cholera-related deaths in 2019, comprising 79% of the global total. Within Africa, the Central African Republic and Nigeria reported the highest mortality rates, at 44.63 and 40.74 per 100,000 of the population, respectively. This concerning rise in cholera mortality has persisted despite global advancements in case management, hygiene, and sanitation, underscoring the enduring challenge of cholera as a global health crisis.

Cholera incidence has been closely linked to climate change. Variations in rainfall patterns, temperature, and the occurrence of floods and drougts have been associated with increased cholera incidence in both African and Asian countries (2). Heavy rainfall can lead to flooding, causing contamination of rivers through the mixing of water with sewer systems. Conversely, low rainfall during droughts reduces access to sufficient safe water, thereby heightening the risk of cholera outbreaks (2). In Africa, countries such as Ghana, Tanzania, and Zambia have demonstrated a positive correlation between rainfall and cholera epidemics (2).

In Malawi, cholera was first reported in 1973 during the seventh cholera pandemic wave caused by *Vibrio cholerae* serotype Inaba, which affected East Africa (3). Since then, cholera has remained endemic in Malawi, with seasonal outbreaks leading to as many as 22,427 cases in a single year (1975) (3). Despite its endemic status, Malawi experienced its deadliest cholera outbreak beginning in March 2022 (4). Where as of March 6, 2023, this outbreak resulted in 51,568 reported cases and 1,612 fatalities (Case Fatality Rate = 3.1%), affecting all districts in the country. Similar to outbreaks in other African nations, this crisis was triggered and exacerbated by severe rainfall events, including Tropical Cyclones Ana, Gombe, and Freddy (3). These cyclones displaced communities and destroyed more than 80 healthcare facilities, further compounding the health crisis (3–5).

Despite systemic improvements in Malawi’s healthcare infrastructure, including experience gained from managing outbreaks of COVID-19 in 2020 and wild poliovirus type 1 in 2022 (6, 7), the 2022 cholera outbreak was the worst in the country’s history. This outbreak recorded a case fatality rate of 3.1%, which is more than the 1% threshold by the WHO (8). However by the start of the outbreak, Malawi had already implemented several preventive measures, including the introduction of the Oral Cholera Vaccine (OCV) in June 2017 (9) and community-wide cholera awareness campaigns. These interventions, alongside improvements in water, sanitation, and hygiene (WASH) services, were expected to mitigate cholera transmission and severity. Yet, the outbreak’s unprecedented impact raises critical questions about the factors contributing to its severity, particularly concerning clinical management.

Since the onset of the cholera outbreak in March 2022, no updated evaluations of patient-level risk factors influencing mortality among hospitalized cholera patients had been conducted. This study addressed this gap by characterizing the patient-level factors and behaviors prevalent in fatal cholera cases and investigating the relationship between these factors and mortality outcomes.

## METHODOLOGY

### Design

This was a multisite study conducted in four Malawian cholera treatment units located in the

District Health Offices at Nkhatabay, Lilongwe, Mangochi and Blantyre Districts. Cholera patient records were reviewed using a case-control methodology, where deceased cholera patients (Cases) were matched with Controls, who were patients also treated for cholera and discharged from the same treatment unit. Data collection took place between 21 August - 1 September 2023 and the duration of file review was between March 2023 – September 2023.

### Study Setting and Eligibility Criteria

High-burden cholera Districts were selected from the Northern, Central, and Southern Regions of Malawi based on national cholera surveillance data from the Malawi Ministry of Health (https://cholera.health.gov.mw/surveillance). The study included patient medical records from all cholera diagnosed patients admitted to the selected treatment units. Eligibility was restricted to patients with a confirmed cholera diagnosis through either a rapid antigen test or a positive cholera stool culture.

### Survey Tool

A mortality audit tool was developed and used for data collection. The tool consisted of four main components:

1. **Socio-demographic characteristics**: Included variables such as sex, age, marital status, residential location, and employment status.
2. **Identified health-seeking behaviors**: Assessed cholera vaccine status and time to hospital presentation after symptom onset.
3. **Risk factors**: Investigated patient nutritional status (measured by Body Mass Index and/or Mid-Upper Arm Circumference) and comorbidities such as Human Immunodeficiency Virus (HIV) infection.
4. **Patient treatment assessment**: Included the initial hydration status on admission, adequacy of fluid management (highlighting any discrepancy between initial hydration status and documented IV fluid treatment in the initial 2-6 hrs that did not align with the standard treatment guideline concerning treating no dehydration, some dehydration and severe dehydration) and antibiotic treatment (whether antibiotics were given or not).

### Data Collection

Data were collected by trained research assistants using a digitized mortality audit tool designed on Kobo Toolbox. All patient information was treated confidentially and securely stored on a password-protected computer accessible only to the study team.

### Sample Size Estimation and Sampling technique

A total of 174 cholera patients were enrolled in this study, with 50% classified as cases (patients who died) and 50% as controls (survivors), maintaining a 1:1 ratio. The sample size was determined to achieve a power of 99.97%, assuming a binomial distribution and applying an arcsine transformation to standardize the effect size (h = 0.81). The power analysis was conducted with a significance level of 0.05 under a two-sided alternative hypothesis, ensuring that the study was adequately powered to detect differences between the two groups.

All mortality cases (patients who died from cholera) were first identified in each cholera unit. Then a systematic sampling was applied, where every 2nd patient file was chosen for enrolment into the study. Controls were matched to cases based on age and district of cholera diagnosis to minimize confounding factors.

### Statistical Analysis

The statistical analysis was designed to address two objectives: (1) to characterize patient-level factors and behaviors prevalent in fatal cholera patients and (2) to investigate the association between patient-level factors and cholera mortality.

The data comprised patient records from cholera treatment units, including variables on demographics, clinical presentation, treatment details, and outcomes. Prior to analysis, the dataset was cleaned to remove unnecessary columns, address missing data, and reformat variables into appropriate categories or binary formats. Cases, defined as deceased patients, were matched to controls, defined as survivors, based on age and District of diagnosis. This matching process ensured comparability between groups, with the final dataset consisting of matched cases and controls.

To explore the first objective, baseline characteristics of cases and controls were summarized descriptively. Frequencies and percentages were calculated for categorical variables and Chi-square tests applied to assess associations between these variables. Summary tables were generated to depict distributions of key characteristics, such as age, hydration status, and fluid management. These analyses provided a foundational understanding of the factors prevalent in fatal cholera cases.

For the second objective, a conditional logistic regression model was employed to identify associations between patient-level factors and mortality outcomes, accounting for the matched study design. The binary response variable distinguished survival status (alive = 0, deceased = 1). Predictor variables included time to hospital presentation, clinical status on arrival, hydration status, adequacy of fluid management, time spent in the ward, and antibiotic administration. Non-significant predictors were removed stepwise using the drop1() function with Chi-square tests. Multicollinearity among predictors was assessed using the Variance Inflation Factor (VIF), and variables with high collinearity (with VIF > 10) were excluded from the model. Odds ratios (ORs) with 95% confidence intervals (CIs) were calculated for all predictors in the final model, and statistical significance was set at p < 0.05. Finally, to evaluate the performance of the final model, Likelihood Ratio Tests (LRT) were conducted to compare it to a null model.

All analyses were performed using R (version 4.3.3) (10) with relevant statistical packages, including tidyverse, gtsummary, and survival. Finally, the **S**trengthening the Reporting of Observational studies in Epidemiology (STROBE) reporting guidelines for case-control studies was used when writing this report (11).

### Ethical Considerations

The study adhered to rigorous ethical standards to ensure the privacy and confidentiality of patient information. All data collected were anonymized, with identifiers removed to protect the identity of the individuals whose medical records were reviewed. As the study was a retrospective review of medical records, obtaining direct patient consent was not necessary. However, institutional approvals were secured through support letters from the respective Research Coordinating

Committees of the participating institutions. Furthermore, ethical approval (Protocol #23/04/4042) for the study was granted by the National Health Sciences Research Committee (NHSRC) in Malawi, ensuring compliance with national ethical guidelines for research.

## RESULTS

### Baseline characteristics

The study analyzed 174 eligible cholera patient files, consisting of 87 identified cases matched to corresponding controls. **Table 1** summarizes the baseline characteristics of cases and controls.

**Table 1:** Baseline characteristics of enrolled cholera patients categorized as cases and controls.

| Variable | N | case, N = 87<sup>1</sup> | control, N = 87<sup>1</sup> | p-value<sup>2</sup> |
| --- | --- | --- | --- | --- |
| **Age range of Participant (years)** | 174 |  |  | >0.9 |
| <10 |  | 17 (20%) | 18 (21%) |  |
| 10-19 |  | 16 (18%) | 15 (17%) |  |
| 20-29 |  | 16 (18%) | 15 (17%) |  |
| 30-39 |  | 14 (16%) | 14 (16%) |  |
| 40-49 |  | 7 (8.0%) | 8 (9.2%) |  |
| 50-59 |  | 6 (6.9%) | 6 (6.9%) |  |
| >60 |  | 11 (13%) | 11 (13%) |  |
| **Gender status** | 174 |  |  | 0.3 |
| Female |  | 37 (43%) | 42 (48%) |  |
| Male |  | 48 (55%) | 45 (52%) |  |
| Unknown |  | 2 (2.3%) | 0 (0%) |  |
| **Marital Status** | 174 |  |  | >0.9 |
| Single |  | 40 (46%) | 40 (46%) |  |
| Married |  | 38 (44%) | 37 (43%) |  |
| Unknown |  | 9 (10%) | 10 (11%) |  |
| **District** | 174 |  |  | 0.070 |
| Central(Lilongwe) |  | 3 (3.4%) | 13 (15%) |  |
| Northern(Nkhatabay) |  | 13 (15%) | 10 (11%) |  |
| Southern (Blantyre) |  | 8 (9.2%) | 8 (9.2%) |  |
| Southern(Mangochi) |  | 63 (72%) | 56 (64%) |  |

<sup>1</sup>n (%)
<sup>2</sup>Pearson's Chi-squared test

Among the enrolled patients, the largest proportion, comprising 20% (n = 17/87) in cases and 21% (n = 18/87) in controls, were children under 10 years old. A majority of patients across both groups were male, making up 55% (n = 48/87) in cases and 52% (n = 45/87) in controls. Additionally, most patients were single, with 46% (n = 40/87) of cases and controls reporting single marital status.

Regarding the geographical distribution, most enrolled patients originated from Mangochi District, accounting for 72% (n = 63/87) in cases and 64% (n = 56/87) in controls. No statistically significant differences were identified for proportions in age range, gender status, marital status , or District between the two groups (p-values > 0.05).

### Objective 1: Patient-level factors in cholera patients

#### Time to hospital presentation after symptom onset

**Fig 1** illustrates the proportion of cholera patients who presented to the hospital at varying intervals after symptom onset, stratified by survival outcome (alive for controls vs. deceased for cases). The time intervals analysed include <6 hours, 6–24 hours, 24–48 hours, and >48 hours.

**Fig 1:**
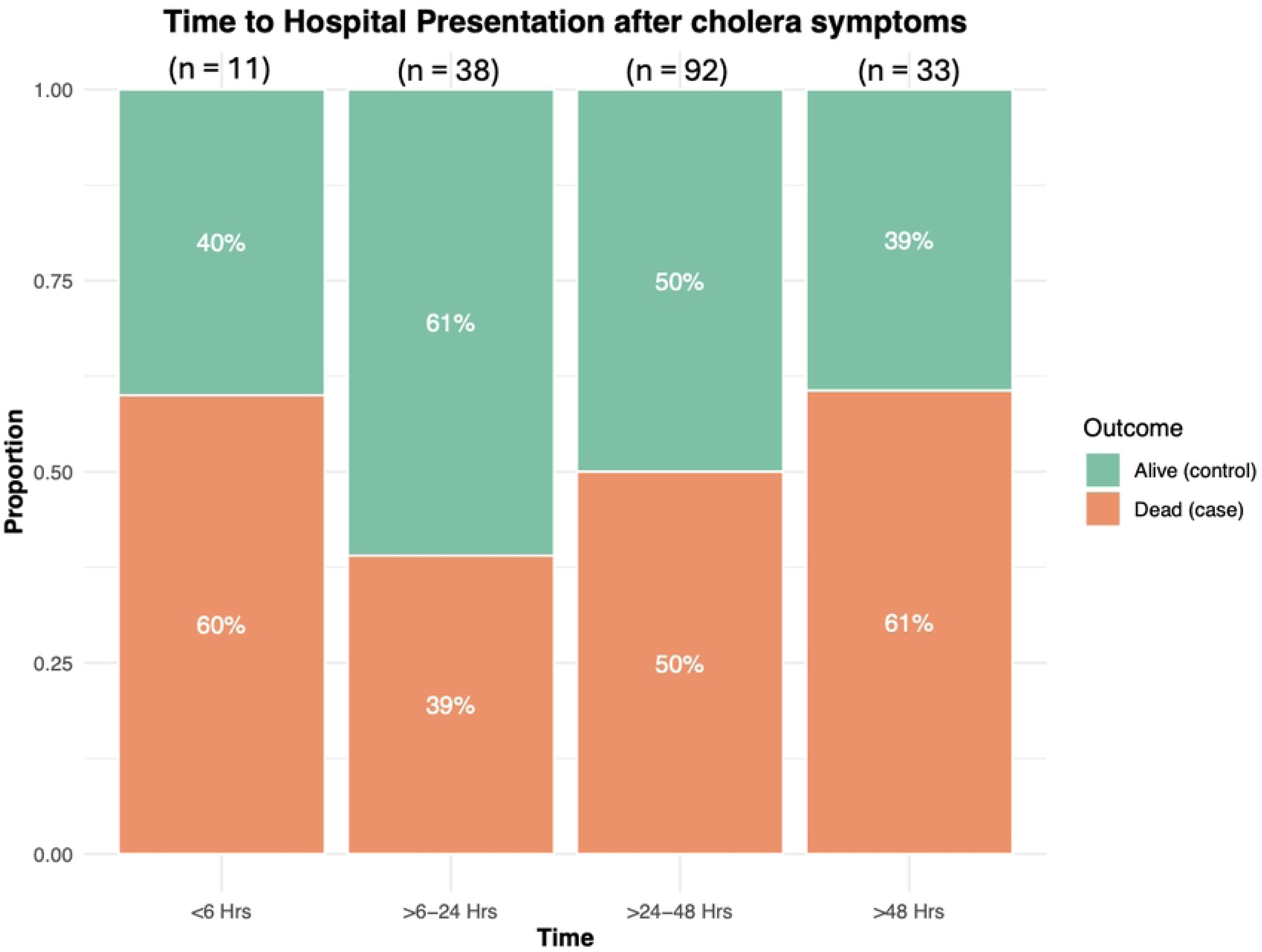
Time to hospital presentation among cholera patients stratified by survival outcome. The bar graph depicts the proportion of cholera patients who presented to the hospital within four time-intervals (<6 hours, 6–24 hours, 24–48 hours, >48 hours) based on their survival outcome (alive vs. deceased).

Among patients who survived (controls), a larger proportion presented within the first 6-24 hours of symptom onset (61%) compared to those who succumbed to the infection (cases), where only 39% presented within this timeframe. Conversely, a greater proportion of deceased patients presented after 48 hours (61%) compared to survivors (39%). No statistically significant differences were identified in the proportions of time to presentation after symptom onset between cases and controls (p-value = 0.3).

#### Reported hydration status on admission

**Fig 2** illustrates the proportions in the hydration status of cholera patients upon hospital admission, categorized as either “well hydrated” , “some dehydration” and “severe dehydration” and stratified by survival outcome (alive vs. deceased).

**Fig 2:**
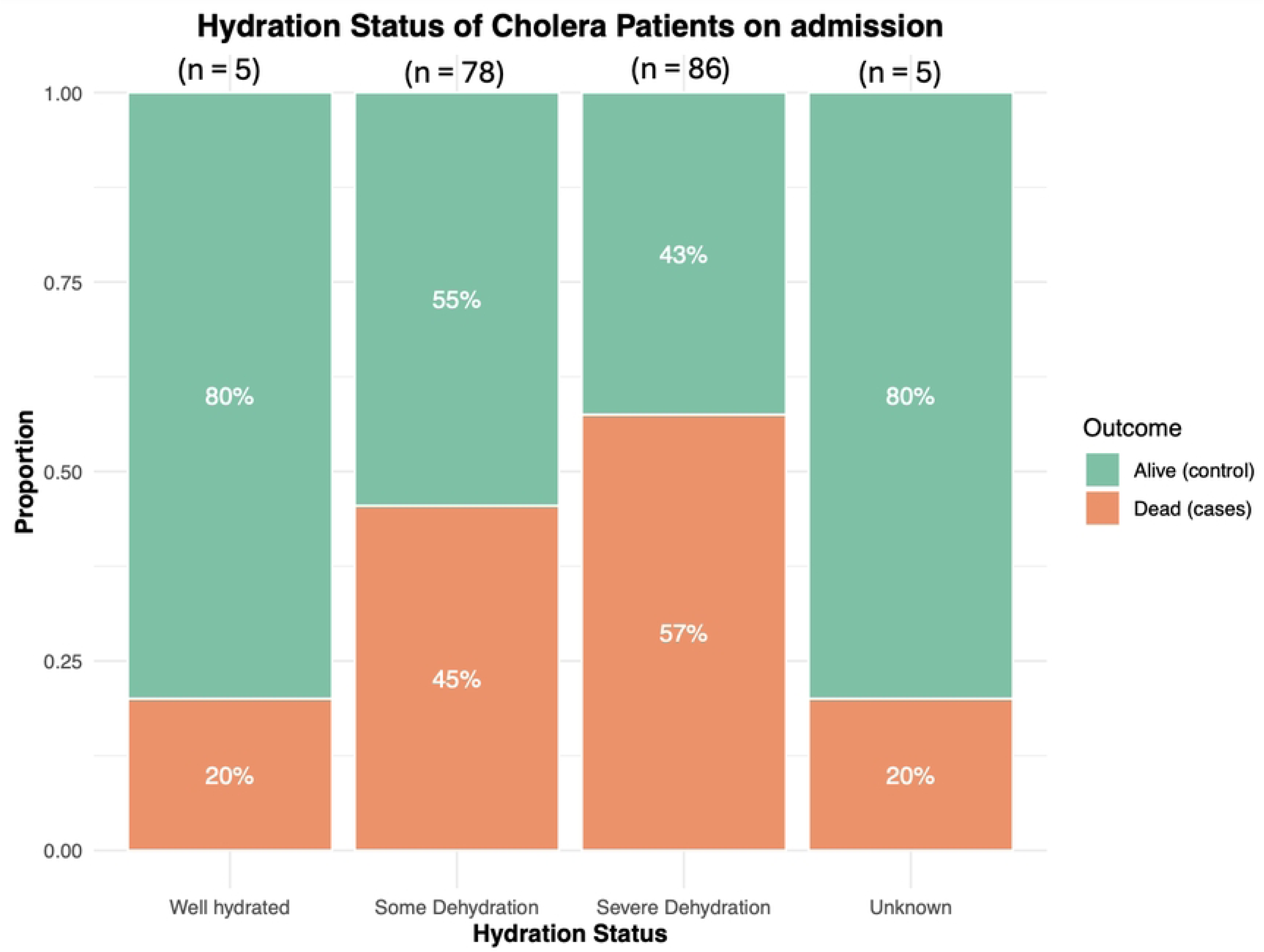
Hydration status of cholera patients on admission stratified by survival outcome. The bar graph depicts the proportion of patients who presented well hydrated, with some dehydration and severely dehydrated as reported after initial clinical assessment.

A higher proportion of survivors (controls) were well hydrated on admisison (80%) compared to deceased patients (cases), of whom only 20% were in well hydrated. Conversely, higher proportions of cases than controls were consistently classified with some dehydration and severely dehydrated on admission, with 53% and 52 % respectively. However, no statistically significant differences were identified in these proportions between cases and controls (p-value = 0.10).

#### Adequacy of fluid management

**Fig 3** highlights the adequacy of intravenous (IV) fluid management among cholera patients, categorized as either adequately or inadequately managed based on their clinical condition at admission and stratified by survival outcome.

**Fig 3:**
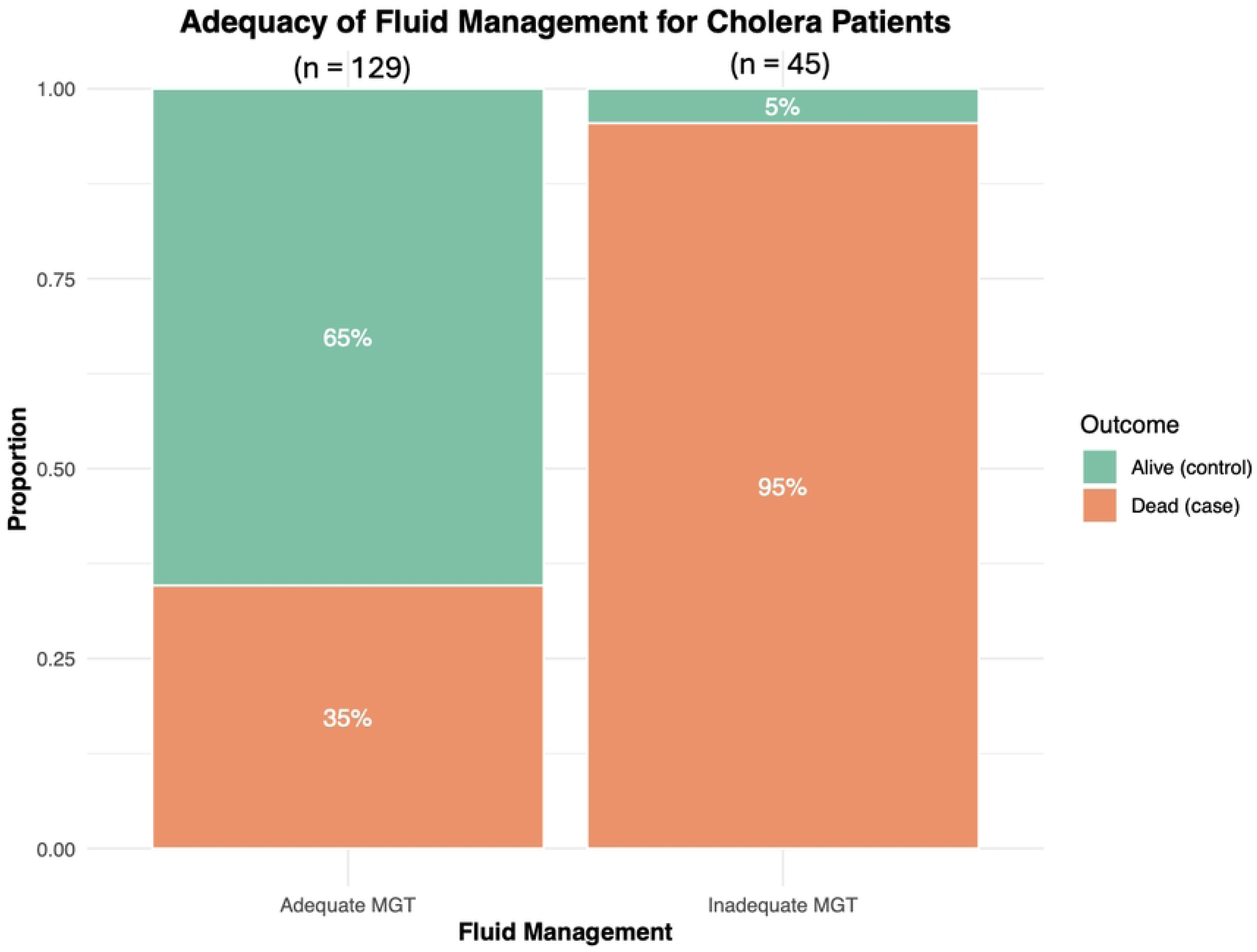
Adequacy of fluid management in cholera patient stratified according to survival outcome. This bar graph depicts fluid management categorized as adequate and inadequate depending on the clinical condition of the patient on arrival to the hospital.

The highest proportion of deceased patients (cases) (95%) were classified as having received inadequate IV fluid management, while only 5% of controls fell into this category. In contrast, a higher proportion of survivors (controls) (65%) were classified as adequately managed with IV fluids compared to 35% of cases. These observed differences in the proportion of fluid management adequacy between cases and controls were statistically significant (p-value < 0.001).

#### Time spent in the hospital ward

**Fig 4** presents the distribution of total time spent in the hospital ward by cholera patients, categorized into four time intervals (6–24 hours, 24–48 hours, 48 hours–7 days, and >7 days) and stratified by survival outcome (deceased vs. alive).

**Figure 4:**
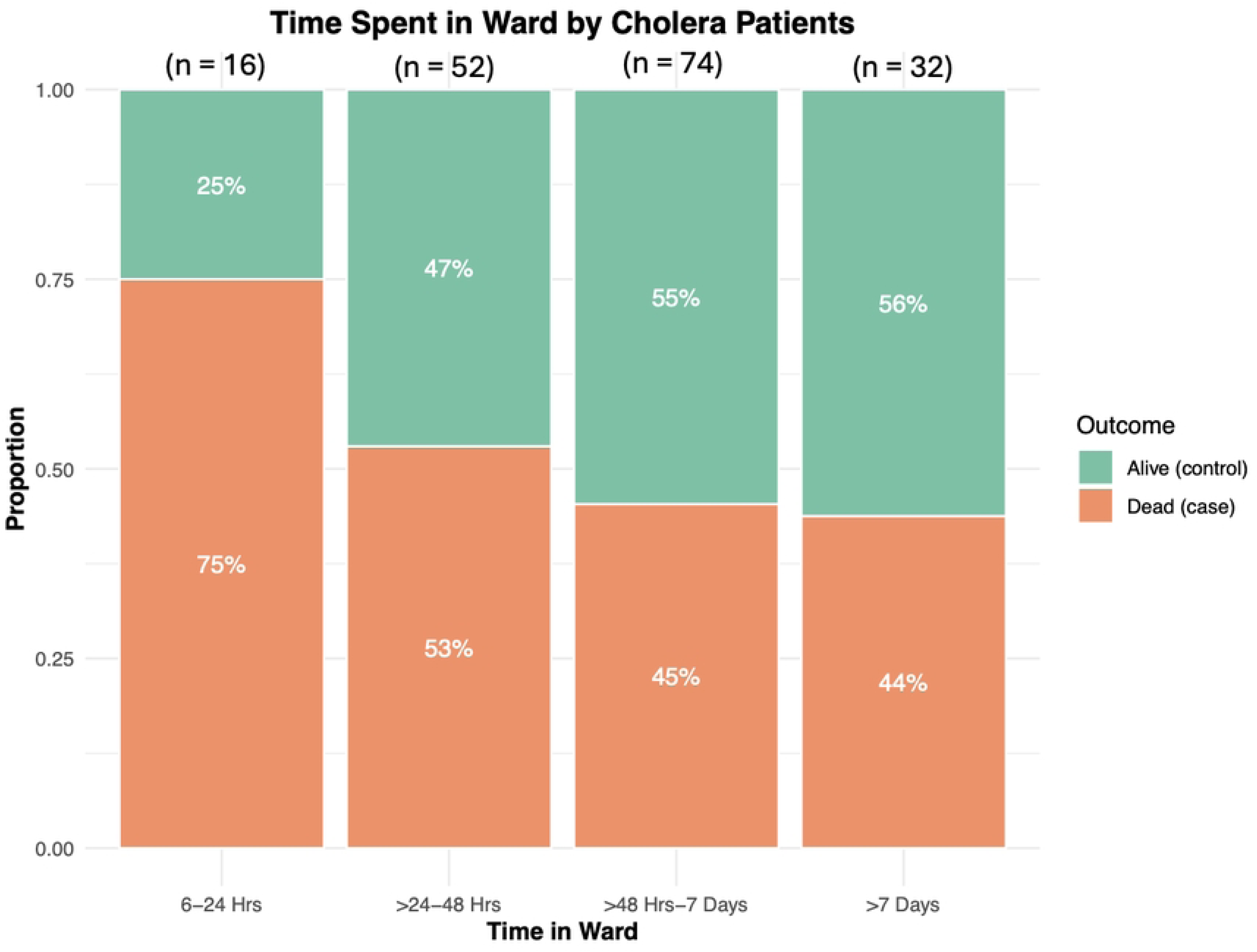
Time spent in the hospital ward by cholera patients, categorized into four time-intervals and stratified by mortality status.

The highest proportion of deceased patients (75%) spent less than 24 hours in the ward, compared to only 25% of survivors (controls) in this time interval. Conversely, the highest proportion of survivors (56%) spent > 7 days hours in the hospital, while 44% of deceased patients spent this duration in the ward. Notably, among patients who stayed longer than 48 hours, survivors outnumbered deceased patients.These differences in the proportions observed between cases and controls were not statistically significant (p-value = 0.15).

### Objective 2: Association between patient-level factors and mortality

**Table 2** presents the association between patient-level factors and mortality among cholera patients, as determined by the final conditional logistic regression model. The final model retained two significant predictors: time to hospital presentation and adequacy of fluid management. Time to presentation was categorized into four intervals: <6 hours, 6–24 hours, 24–48 hours, and >48 hours, while adequacy of fluid management was classified as adequate or inadequate. This model demonstrated a significant ability to explain variance in the data (LRT p < 0.001).

**Table 2:** Conditional logistics regression model depicting factors associated with cholera mortality.

| Characteristic | OR<sup>1</sup> | 95% CI<sup>1</sup> | p-value |
| --- | --- | --- | --- |
| **Time from symptom onset to hospital presentation** |  |  |  |
| <6 Hrs | — | — |  |
| >6-24 Hrs | 0.34 | 0.05, 2.12 | 0.2 |
| >24-48 Hrs | 0.70 | 0.13, 3.70 | 0.7 |
| >48 Hrs | 1.58 | 0.26, 9.73 | 0.6 |
| **Fluid Management** |  |  |  |
| Adequate MGT | — | — |  |
| Inadequate MGT | 117 | 14.3, 959 | <0.001 |

<sup>1</sup>OR = Odds Ratio, CI = Confidence Interval

The stepwise selection process using “drop1” indicated that fluid management had the highest likelihood ratio test value (LRT = 50.337, p < 0.001). Time to presentation was also a significant predictor (LRT = 9.064, p = 0.02845). Other variables, such as marital status, status on arrival, hydration status, antibiotic management, and time spent in the ward, were not statistically significant and were excluded from the final model (p-values > 0.05). HIV status and vaccine status were removed from the model because they contained many missing values. Finally, the inclusion of gender as a variable caused the conditional logistic regression model to fail to converge. To ensure the stability and reliability of the model, gender was excluded from the final analysis. No multicollinearity was detected in the final model.

The analysis revealed that patients who presented to the hospital more than six hours after symptom onset had varied odds of mortality compared to those who presented within six hours. Specifically, those presenting within 6–24 and 24 -48 hours had lower odds of mortality (OR = 0.34, 95% CI: 0.05–2.12, p = 0.2; OR = 0.70, 95% CI: 0.13–3.70, p = 0.7, respectively), while those presenting after 48 hours had higher odds of mortality (OR = 1.58, 95% CI: 0.26–9.73, p = 0.6). However, none of these associations were statistically significant.

In contrast, inadequate fluid management was strongly associated with increased odds of mortality, with an odds ratio of 117 (95% CI: 14.3–959, p < 0.001).

## DISCUSSION

This case-control study characterizes patient-level factors and clinical management practices associated with mortality among cholera patients in Malawi’s treatment units. The findings reveal that inadequate intravenous fluid management is strongly associated with fatal outcomes, significantly increasing the odds of mortality by over 100-fold compared to patients who received adequate IV fluid management. This underscores the critical role of prompt and appropriate fluid resuscitation in the management of cholera, particularly for patients presenting with dehydration.

Among the fatal cholera cases in this study, 95% were classified as having received inadequate intravenous fluid management, highlighting a critical gap in adherence to established treatment guidelines. The cholera toxin’s action on the intestinal mucosa leads to profound fluid loss, resulting in hypovolemic shock and metabolic acidosis (12). This underscores the importance of appropriate and timely fluid resuscitation in managing cholera patients. According to the World Health Organization’s recommended treatment for severe cholera, patients require rapid IV rehydration within the first 2–6 hours, followed by antibiotics and Oral Rehydration Salts (ORS) over the next two days (8). On average, severe cases necessitate approximately 7 litres of intravenous fluids and 14 litres of ORS over the course of treatment (12). Given these significant fluid requirements, any deviation or delay in administering adequate rehydration therapy can have severe and often fatal consequences, emphasizing the pivotal role of strict adherence to fluid management protocols in reducing mortality among cholera patients.

Our findings on the strong association between inadequate IV fluid management and cholera mortality are consistent with existing literature (13) . For instance, during one of Congo’s worst cholera outbreaks in 1994, where an estimated 12,000 refugees died, inadequate fluid management was attributed to a slow rate of IV fluid administration or improper use that did not align with standard treatment guidelines (13). These deficiencies were largely due to shortages of IV fluids or water, the latter impacting the availability of Oral Rehydration Salts (13). Similarly, in this study, the inadequate availability of IV fluids in Malawian treatment units was attributed to resource constraints, as previously documented (4, 5). This was exacerbated by infrastructural damage to water and sanitation systems and the overcrowding caused by displacement from Tropical Cyclone Freddy. These factors overwhelmed the healthcare system, leaving it unable to meet the average IV fluid requirement of approximately 7 litres per cholera patient within the critical first 2–6 hours of care. To address such challenges, we emphasize the importance of strengthening cholera surveillance systems in Malawi by integrating meteorological data to predict climate-related influences on health and improve preparedness for future outbreaks.

The study further demonstrated the potential influence that dehydration severity on admission can have on patient outcomes. We observed that patients presenting with severe dehydration were overrepresented among fatal cases, indicating that delays in initial resuscitation and fluid replacement likely contributed to the observed mortality. While hydration status on admission did not yield statistically significant associations with mortality in this study, the trend observed aligns with existing evidence from cholera outbreaks in other African countries, where severe dehydration, alongside older age, gender and residential location, are key predictors of poor outcomes (14). Therefore, early identification and aggressive management of dehydration in cholera patients remain pivotal in reducing fatalities.

Time to hospital presentation was also assessed as a potential factor influencing mortality. Although the findings in this study were not statistically significant, the trends suggested that delayed presentation beyond 48 hours after symptom onset increased the likelihood of mortality. This delay likely reflects the existing barriers to healthcare access in Malawi, as highlighted by Msokwa (15), including geographic challenges, financial constraints, and infrastructural limitations. Such barriers can exacerbate cholera outcomes during outbreaks. Consistent with this observation, Kelly Osezele Elimian, Anwar Musah (16) demonstrated that a delay of more than two days after the onset of cholera symptoms significantly increased the odds of cholera-related death (aOR 2.08, 95% CI: 1.52–2.85). These findings reinforce the critical need for strengthening community-based awareness campaigns and implementing robust early referral systems to ensure timely access to care, thereby improving patient outcomes during cholera outbreaks.

While this study provides valuable insights into the factors associated with cholera mortality in Malawi, several limitations must be acknowledged. The retrospective design and reliance on medical record reviews may have introduced information bias, particularly in the categorization of fluid management adequacy. This categorization depended on the quality of documentation, and any errors or inconsistencies in recording the volume, type, and timing of IV fluids administered could have resulted in the misclassification of patients. Such misclassification may have distorted the true relationship between fluid management and mortality. However, this limitation was mitigated by employing trained medical officers to lead data collection teams, ensuring that patients were carefully categorized as “adequately” or “inadequately” managed with IV fluids based on thorough reviews of patient files.

Additionally, the lack of granular data on key patient-level factors, such as health-seeking behaviors, may have reduced the statistical power to detect significant associations for some predictors. The absence of detailed data on health-seeking behaviors, for instance, limits the ability to identify root causes of delayed hospital presentations and broader social or economic barriers to healthcare access. Furthermore, variables such as cholera vaccination status and HIV status were excluded from the final model due to substantial missing data, attributed to incomplete patient file documentation. These factors compromise the generalisability of the findings to all Malawian treatment units.

Despite these limitations, the findings highlight critical gaps in cholera treatment and provide a basis for targeted interventions. Future studies should aim to address these challenges by employing prospective designs and utilizing comprehensive data collection tools to minimize information bias and capture detailed patient-level factors. These efforts will improve the ability to identify actionable strategies for reducing cholera mortality.

## CONCLUSION

This study highlights the critical role of adequate intravenous fluid management in reducing cholera mortality, underscoring significant gaps in adherence to treatment protocols within Malawian treatment units during the 2022 outbreak. Delays in hospital presentation and resource constraints further compounded the challenges, highlighting systemic barriers to effective care delivery during outbreaks. Strengthening cholera surveillance systems and implementing robust community-based awareness and referral systems are essential to improving outbreak preparedness and patient outcomes. Addressing these gaps through targeted interventions and future prospective studies will be pivotal in mitigating cholera-related mortality in Malawi and similar settings.

## Data Availability

Data cannot be shared publicly because the data is owned by the Malawi Ministry of Health (MOH). The authors were only given permission to use the patient files and data gathered for the purposes of the request submitted to the MOH.

## ACKNOWLEDGEMENTS

This case-control study was made possible because of the financial support provided by the Research department at the Public Health Institute of Malawi. We extend our deepest gratitude to the District Health Offices at Nkhatabay, Lilongwe, Mangochi and Blantyre Districts for their endorsement to implement this work.

## Notes

### Competing Interest Statement

The authors have declared no competing interest.

### Funding Statement

The authors did not receive any funding for this work.

### Author Declarations

Ethical approval (Protocol #23/04/4042) for the study was granted by the National Health Sciences Research Committee (NHSRC) in Malawi, ensuring compliance with national ethical guidelines for research.

